# Sex differences in perihematomal edema volume and outcome after intracerebral hemorrhage

**DOI:** 10.1101/2023.09.28.23296302

**Authors:** Jens Witsch, Quy Cao, Jae W. Song, Yunshi Luo, Kelly L. Sloane, Aaron Rothstein, Christopher G. Favilla, Brett L. Cucchiara, Scott E. Kasner, Steve R. Messé, Huimahn A. Choi, Louise D. McCullough, Stephan A. Mayer, Aaron M. Gusdon

## Abstract

**Objective:** To determine whether in patients with intracerebral hemorrhage (ICH) perihematomal edema (PHE) volume trajectories differ by sex.

**Methods:** We conducted a post-hoc analysis of the Factor-VII-for-Acute-Hemorrhagic-Stroke-Treatment (FAST) trial that randomized patients with ICH to receive recombinant activated Factor VIIa or placebo. Computerized planimetry calculated PHE and ICH volumes on serial CT scans (at baseline [within 3 hours of onset], at 24, and at 72 hours). Generalized estimating equations examined interactions between sex, CT-timepoints, and FAST treatment-arm on PHE and ICH volumes. Mixed and multivariate logistic models examined associations between sex, PHE, and outcomes.

**Results:** 781 with supratentorial ICH (mean age 65 years) were included. Compared to women (n=296), men (n=485) had similar median ICH (14.9 versus 13.6 ml, p=0.053), and PHE volumes (11.1 versus 10.5 ml, p=0.56) at baseline but larger ICH and PHE at 24 hours (19.0 versus 14.0, p<0.001; 22.2 versus 15.7, p<0.001) and 72 hours (16.0 versus 11.8, p<0.001; 28.7 versus 19.9, p<0.001). Men had higher absolute PHE expansion (p<0.001), and more hematoma expansion (growth ≥33% or 6 mL at 24 hours, 33% versus 22%, p<0.001). An interaction between sex and CT-timepoints on PHE (p<0.001) but not on ICH volumes confirmed a steeper PHE trajectory in men. PHE expansion (per 5mL, odds radio, 1.19, 95%-confidence interval 1.10-1.28), but not sex, was associated with poor outcome.

**Conclusions:** PHE expansion and trajectory in men were significantly higher. PHE expansion was associated with poor outcomes independent of sex. Mechanisms leading to sex differences in PHE trajectories merit further investigation.

**What is already known on this topic:** Prior research has reported sex differences in intracerebral hemorrhage (ICH) characteristics and some studies suggest worse outcome after ICH in women. However, we do not have a good understanding whether there are sex differences in perihematomal edema (PHE) volume trajectories, or whether sex, independent of confounders, is associated with poor after ICH.

**What this study adds:** In this post-hoc analysis of 781 patients with supratentorial ICH from the Factor-VII-for-Acute-Hemorrhagic-Stroke-Treatment (FAST) trial in which patients underwent brain CT imaging time-locked to symptom onset (within 3 hours of symptom onset, at 24 hours, and at 72 hours), men compared to women had similar ICH and PHE volumes at baseline, but larger ICH expansion and PHE expansion on follow up imaging. The PHE but not the ICH volume trajectory across scans was significantly higher in men than in women. While PHE expansion was associated with poor outcome at 90 days, outcome between the sexes was similar at 90 days, and sex was not associated with outcome.

**How this study might affect research, practice or policy:** The finding of heightened early PHE and ICH expansion in men may inform study design, patient recruitment strategies, and pre-specification of subgroup analyses in future interventional trials. The findings of this study also suggest that focusing on sex-specific factors may allow novel mechanistic insight into PHE, a major cause of secondary injury and poor outcome after ICH.

## INTRODUCTION

Intracerebral hemorrhage (ICH) accounts for 10-15% of all strokes and results in high rates of morbidity and mortality, but effective therapies have remained elusive.^1^ ^2^ Intraparenchymal hematoma directly causes tissue injury, and hematoma expansion has been consistently linked to worse outcomes.^3^ In addition to this primary injury, secondary injury occurs, of which perihematomal edema (PHE) is a radiographic surrogate.^4^ ^5^ Although early PHE expansion has been associated with poor outcomes, controversy remains regarding the relationship between PHE and outcome.^6–11^ Prior studies have been limited by variable timing of serial neuroimaging, which confounds the association between PHE and outcome.

Superimposed onto the unanswered question whether PHE is associated with poor outcome, is the question whether PHE volumes and trajectories segregate by sex. Increasing efforts to characterize sex differences in ICH characteristics have led to the findings of differences in ICH location (greater frequency of deep ICH in men) and ICH volume (larger ICH volume in men), and there have been suggestions of higher rates of hematoma expansion in men.^2^ ^12–15^ However, the effect of sex on PHE trajectories has not been well characterized.

Herein, we determined 1) the effect of sex on PHE and ICH, and 2) the association between PHE and 90-day outcome using data from the Recombinant Factor VIIa in Acute Intracerebral Hemorrhage (FAST) trial with serial neuroimaging performed at standardized time points.

We hypothesized that men would have larger PHE and ICH volumes than women, which would result in worse outcomes in men. We further hypothesized that PHE volume and PHE expansion would have an independent association with poor outcomes in both sexes.

## METHODS

Additional descriptions of the methods used in this study are available in the supplementary material methods.

### Design and participants

We performed a secondary observational (post-hoc) cohort analysis using data from the FAST trial, a multicenter, randomized, double-blind, placebo-controlled trial of recombinant activated Factor VIIa (rFVIIa) for the treatment of spontaneous ICH that randomized 841 patients to receive one of the three treatments within 4 hours of stroke onset: placebo, 20 μg rFVIIa/kilogram body weight, or 80 μg rFVIIa/kilogram body weight.^16^ In this analysis, we excluded patients with infratentorial hemorrhage (n=40), due to the limitations of CT assessment of PHE in the posterior fossa (supplemental figure 1). This retrospective study was exempt from additional, local IRB approval. This study followed the STROBE guideline.

### Neuroimaging

Patients enrolled in the FAST trial underwent CT within 3 hours of symptom onset and at 24 (range: 21-27) and 72 (range: 66-78) hours after administration of the investigational product. Parenchymal and intraventricular hemorrhage (IVH) volumes and PHE volumes were calculated semi-automatically using computerized planimetry (Analyze software, Overland Park, KS) by two neuroradiologists who were blinded to treatment assignment.^16^ In addition to PHE volumes, we report ICH and total lesion volumes, the latter being the sum of PHE, ICH, and IVH volumes (in those patients with an IVH component), as previously reported.^16^ We defined hematoma expansion (HE) as growth in ICH volume by ≥33% or ≥6 mL from baseline to 24 hours.^3^ ^17^

### Functional Outcome

Outcome was assessed at 90 days using the modified Rankin Scale (mRS) in person or through standardized telephone interviews. Poor outcome was defined as mRS 4-6.

### Statistical Analysis

Lesion volumes were measured over time for each subject and were therefore treated as longitudinal data. Because lesion volumes were not normally distributed, we chose generalized Estimating Equations (GEE) models with gamma log-linked generalized linear models that were fitted for PHE, ICH, and total lesion volumes to examine interactions between sex and CT timepoints, as well as between sex and FAST treatment arm assignment. Lesion volumes were naturally log transformed before the analysis. The choice of fitting the data to a gamma distribution was made based on visual inspection. The theoretical densities, cumulative distribution function, and the Q-Q and P-P plots of PHE, ICH, and total lesions volume data are shown in supplemental figures 2-4.

Logistic mixed models tested for associations between PHE volume and poor 90-day outcome accounting for CT timepoint and the following confounders: baseline ICH volume, sex, age, admission Glasgow Coma Scale (GCS) score, ICH location (lobar or non-lobar), presence of IVH, disability prior to the index ICH (pre-morbid mRS), and FAST treatment assignment (placebo, low dose or high dose FVIIa treatment). ICH and PHE volumes were highly correlated (regression coefficients >0.8 at baseline, 24 h and 72 h measurements) and could not be fit within the same model. ICH and PHE volumes were therefore added to two separate outcome models.

We used two strategies to determine the association between PHE volume and outcomes while accounting for the effect of ICH volume and the collinearity between PHE and ICH. First, we calculated the absolute difference (ΔPHE) between PHE volumes (24-h minus baseline volumes and 72-h minus baseline volumes) as measures of PHE expansion and added them to a multivariable logistic model with baseline ICH volume added as a covariate. These models also included a variable accounting for the interaction between PHE expansion and sex. Although the change in PHE between the two time intervals are correlated and do not truly represent two independent models, we chose to report them because both early and later PHE expansion are of clinical interest. Secondly, we regressed PHE volumes on ICH volumes in a linear regression and added the resulting residuals as a covariate to the logistic mixed model of the outcome. The residuals can be interpreted as the influence of PHE volumes on outcome while controlling for ICH volumes.

## RESULTS

### Cohort Characteristics and Lesion Volumes

Of 842 participants of the FAST trial, those with supratentorial ICH (n= 781, mean age 65 years) were included in our analysis (supplemental figure 1). Among these, men (n=485, 62%) and women (n=296, 38%) had a similar prevalence of vascular risk factors (table 1).

**Table 1.** Baseline Characteristics of Patients with Supratentorial Intracerebral Hemorrhage.

|  |  |  |  |
| --- | --- | --- | --- |
| **Characteristic** | **Men**<br>**N = 485** | **Women**<br>**N= 296** | **P**<br>**value** |
| Age, y, mean (SD) | 63.8 (12.5) | 66.4 (13.3) | 0.13 |
| Hypertension | 400 (82.5) | 255 (86.1) | 0.22 |
| Hyperlipidemia | 134 (27.6) | 80 (27.0) | 0.93 |
| Diabetes mellitus | 56 (11.5) | 31 (10.5) | 0.85 |
| Coronary artery disease | 29 (6.0) | 11 (3.7) | 0.35 |
| Atrial Fibrillation | 19 (3.9) | 9 (3.0) | 0.77 |
| Prior ischemic stroke or TIA | 25 (5.2) | 11 (3.7) | 0.70 |
| Baseline mRS |  |  |  |
| 0 | 418 (86.2) | 241 (81.4) | 0.012 |
| 1 | 50 (10.3) | 30 (10.1) |  |
| 2 | 17 (3.5) | 25 (8.4) |  |
| GCS on admission, median (quartiles) | 14 (13-15) | 14 (12-15) | 0.33 |
| Systolic blood pressure on admission, mmHg (SD) | 179 (30) | 183 (30) | 0.054 |
| Treatment assignment |  |  |  |
| 20 µg | 155 (32.0) | 102 (34.5) | 0.51 |
| 40 µg | 170 (35.1) | 108 (36.5) |  |
| Placebo | 160 (33.0) | 86 (29.1) |  |
| Time from symptoms onset to investigational product, minutes (SD) | 159 (36) | 161 (38) | 0.75 |
| Abbreviations: SD, standard deviation; mRS, modified Rankin Scale; GCS, Glasgow Coma scale |  |  |  |
| Data are represented as number (%) unless otherwise stated. |  |  |  |

Women had higher baseline mRS scores than men before the index ICH (p=0.02). Mean [SD] systolic blood pressures between admission and hospital day 3 were similar in men and women (supplemental table 1).

In the overall cohort, ICH volumes increased from a median [IQR] of 14.5 mL [6.9-33.5] at baseline to 16.5 mL [7.4-37.8] at 24-hours (p<0.001), followed by a reduction to a median volume of 14.2 mL [6.6-32.3] at 72-hours (comparing 24 and 72 hours, p<0.001). Median PHE was 11.0 mL [4.9-23.6] at baseline, 19.5 mL [9.5-39.1] at 24 hours, and 24.4 mL [12.1-50.3] at 72 hours (baseline versus 24 hours, p<0.001; and 24-hours versus 72 hours, p<0.001). The median [IQR] total lesion volume was 31.2 ml [15.0-60.7] at baseline, 41.1 ml [20.3-84.9] at 24-hours, and 44.2 ml [22.0-86.6] at 72-hours (both comparisons, p<0.001). A component of intraventricular hemorrhage (IVH) on admission CT was present in 37%.

### Lesion Volumes in Men and Women

Men compared to women had similar median ICH, PHE, and total lesion volumes at baseline (table 2, figure 1).

**Figure 1.**
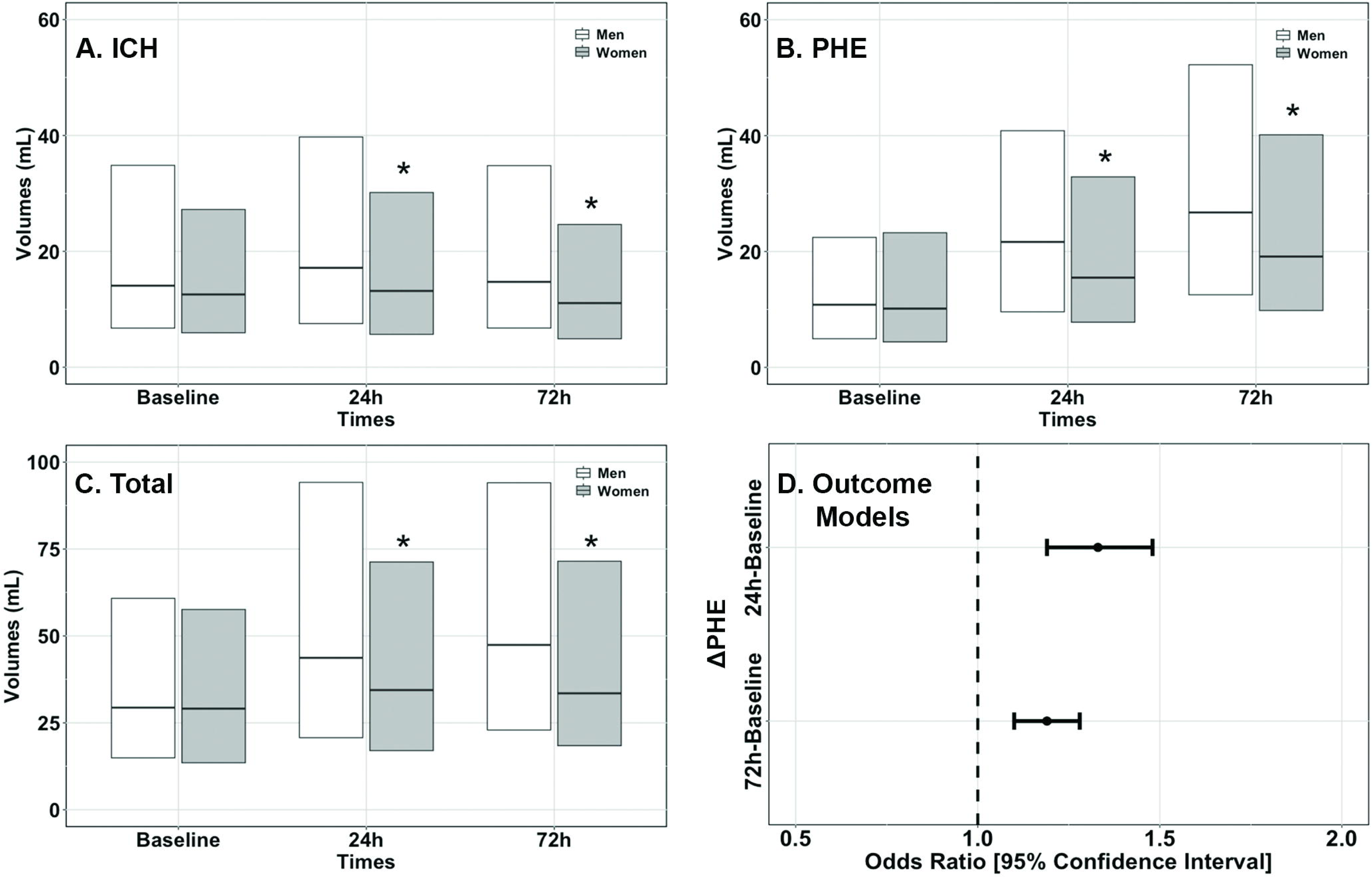
Lesion volumes in men and women with ICH, and associations between PHE expansion and functional outcome. Intracerebral hemorrhage (ICH, **A**), perihematomal edema (PHE, **B**), and total lesion (ICH, PHE, and intraventricular hemorrhage, **C**) volumes in FAST patients with supratentorial ICH (n=781). Depicted are medians and interquartile ranges. Asterisks indicate statistical significance (p<0.05). **D**) Multivariate logistic regression model of PHE expansion between baseline and 24 h and 72 h respectively, adjusted for confounders (listed in Methods). Poor outcome: modified Rankin scale 4-6 at 90 days.

**Table 2.** Imaging Characteristics of Patients with Supratentorial Intracerebral Hemorrhage.

| Characteristic | Men<br>N = 485 | Women<br>N= 296 | P<br>value |
| --- | --- | --- | --- |
| ICH volume, baseline, mL | 14.9 (7.4-36.5) | 13.6 (6.1-29.3) | 0.053 |
| ICH volume, 24 h, mL | 19.0 (8.1-42.5) | 14.0 (6.0-31.5) | <0.001 |
| ICH volume, 72 h, mL | 16.0 (7.3-36.0) | 11.8 (5.2-25.3) | <0.001 |
| PHE volume, baseline, mL | 11.1 (5.1-23.4) | 10.5 (4.4-23.9) | 0.56 |
| PHE volume, 24 h, mL | 22.2 (10.1-42.3) | 15.7 (8.4-34.5) | <0.001 |
| PHE volume, 72 h, mL | 28.7 (13.5-52.9) | 19.9 (10.1-41.3) | <0.001 |
| Total lesion volume, baseline, mL | 31.4 (15.5-61.9) | 30.4 (13.8-58.5) | 0.29 |
| Total lesion volume, at 24 h, mL | 44.8 (22.6-95.5) | 35.3 (17.1-72.3) | <0.001 |
| Total lesion volume, at 72 h, mL | 49.8 (24.4-96.0) | 34.9 (18.9-76.9) | <0.001 |
| Presence of IVH on admission<sup>a</sup> | 170 (35.1) | 121 (40.9) | 0.10 |
| ICH location |  |  |  |
| Lobar<sup>a</sup> | 69 (14.2) | 44 (14.9) | 0.87 |
| Non-lobar<sup>a</sup> | 396 (81.6) | 244 (82.4) |  |
| Hematoma expansion<sup>a, b</sup> | 160 (33.0) | 64 (21.6) | <0.001 |

Abbreviations: ICH, intracerebral hemorrhage; IVH, intraventricular hemorrhage; mL, milliliters; PHE, perihematoma edema.
Data shown as median (interquartile range) unless stated otherwise
<sup>a</sup> Data are represented as number (%)
<sup>b</sup> defined as increase in ICH volume between admission and 24 hours CT of either $\geq 33\%$ or $\geq 6$ ml

The median [IQR] change in PHE volumes between admission and the 24-hour CT was 8.0 mL [2.6-18.0] in men and 4.2 mL [1.5-11.5] in women (between group comparison p<0.001), and between 24-hour and 72-hour CT the change in PHE volume was 6.4 mL [1.9-12.7] in men and 4.0 mL [1.0-8.9] in women (p<0.001).

### PHE and Sex

In GEE models, we found significant effects of the CT timepoints on all lesion volumes (p<0.001 in the ICH, PHE, and total lesion models) indicating that all lesion volumes changed across CT scan timepoints (table 3).

**Table 3.**
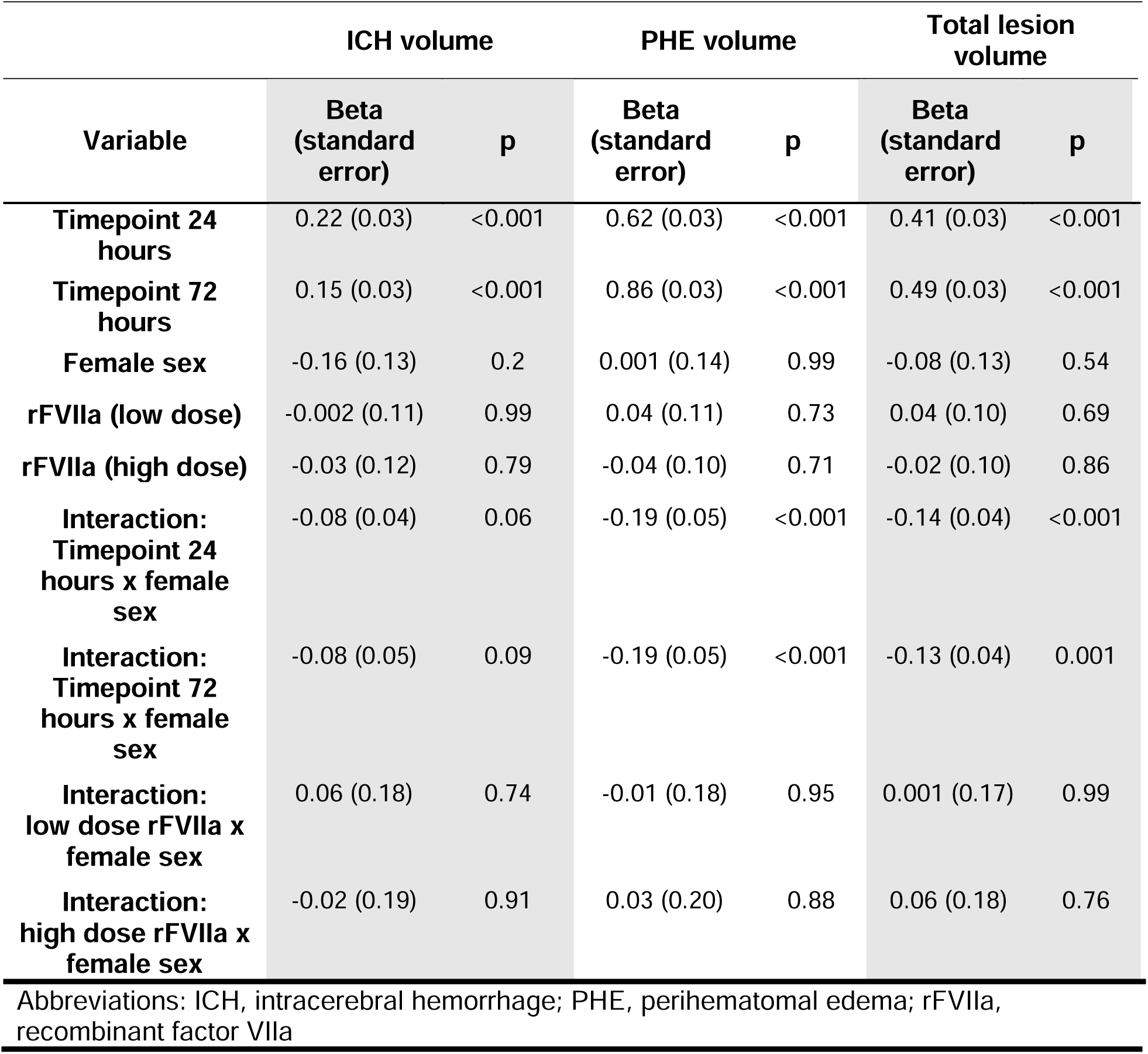
Effects on Lesion Volumes Modelled by Generalized Estimating Equations.

The PHE volume trajectory differed by sex with a higher trajectory in men compared to women, as demonstrated by a significant interaction between sex and CT timepoint (24 hour CT, beta −0.19, standard error, SE, 0.05, p<0.001; 72 hour CT, beta −0.19, SE, 0.05, p<0.001).

Interactions between sex and CT time points were also found for total lesion volume but not for ICH volume (table 3). The interactions between sex and FAST treatment groups did not affect any of the three lesion volumes (all interactions not statistically significant, table 3).

### PHE, Sex, and Functional Outcome

Median mRS at 90 days of the analytical cohort was 3 (IQR 2-5), and 373 (48%) had poor outcome. The median mRS was identical in men and women (median 3, IQR 2-5). 47.6% of men, and 48% of women had poor 90-day outcome (figure 2).

**Figure 2.**
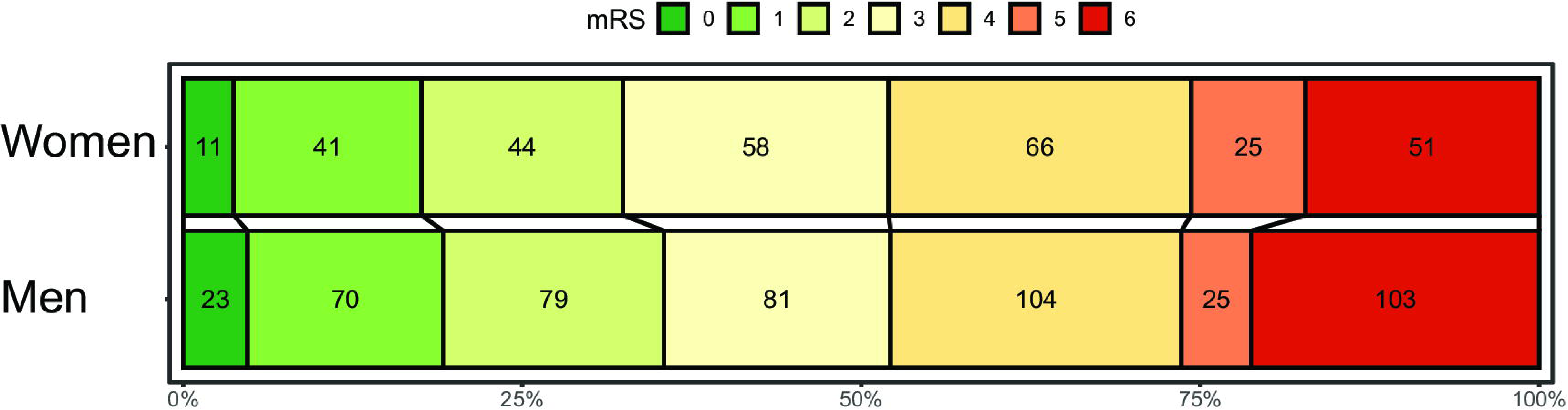
Outcome at 90 days (modified Rankin score, mRS) after spontaneous intracerebral hemorrhage in women and men in the analytical cohort. Numbers indicate n participants per mRS grade 0-6 (color coded). X-axis indicates distribution of mRS score in %.

In separate logistic mixed models testing for associations with poor 90-day outcome, each adjusted for known confounders, both ICH and PHE were associated with poor outcome (supplemental table 3). A multivariate logistic regression model, adjusted for baseline ICH volume, demonstrated that a 5 mL increase in PHE between baseline and 24 hours was associated with a 33% increase in the odds of poor outcome (OR 1.33, CI 1.19-1.48), and a 5 mL increase in PHE between baseline and 72 hours was associated with a 19% increase in the odds of poor outcome (OR 1.19, CI 1.10-1.28). Neither sex nor the interaction between sex and PHE expansion was associated with poor outcome in these models (table 4, figure 1D).

**Table 4.** Multivariate Logistic Regression Model of Poor Outcome (mRS 4-6) at 90 Days.

| Variable | PHE expansion between baseline and 24 hours† |  | PHE expansion between baseline and 72 hours‡ |  |
| --- | --- | --- | --- | --- |
|  | OR (CI) | p | OR (CI) | p |
| **PHE Delta** | 1.33<br>(1.19-1.48) | <0.001 | 1.19<br>(1.10-1.28) | <0.001 |
| **Female sex** | 1.18<br>(0.72-1.93) | 0.52 | 1.10<br>(0.62-1.94) | 0.75 |
| **Interaction: sex\*PHE delta** | 0.99<br>(0.96-1.02) | 0.51 | 1.00<br>(0.97-1.03) | 0.98 |
| **ICH volume at baseline** | 1.32<br>(1.19-1.42) | <0.001 | 1.29<br>(1.20-1.40) | <0.001 |
| **Lobar hematoma location** | 0.25<br>(0.13-0.49) | <0.001 | 0.23<br>(0.11-0.46) | <0.001 |
| **IVH** | 2.10<br>(1.41-3.15) | <0.001 | 2.18<br>(1.45-3.29) | <0.001 |
| **Age** | 1.05<br>(1.04-1.07) | <0.001 | 1.06<br>(1.04-1.08) | <0.001 |
| **GCS on admission** | 0.83<br>(0.75-0.92) | <0.001 | 0.84<br>(0.76-0.93) | 0.001 |
| **mRS prior to the index ICH** | 2.55<br>(1.68-3.88) | <0.001 | 2.31<br>(1.51-3.54) | <0.001 |
| **FAST treatment assignment** | 1.15<br>(0.91-1.44) | 0.25 | 1.23<br>(0.97-1.56) | 0.08 |
| **Time from stroke to treatment#** | 1.00<br>(1.0-1.01) | 0.81 | 1.00<br>(1.00-1.01) | 0.99 |

Abbreviations: OR, odds ratio; CI, 95% confidence interval; GCS, Glasgow Coma Scale; ICH, intracerebral hemorrhage; IVH, intraventricular hemorrhage; PHE, perihematoma edema; rFVIIa, recombinant factor VIIa; mRS, modified Rankin Scale.
†Cases with complete data included in this model: n=708 (91% of cohort)
‡Cases with complete data included in this model: n=670 (86% of cohort)
\*OR are represented as per 5 mL increase in delta PHE or ICH volume.
#Administration of the FAST investigational product

These results were further confirmed by a second analysis accounting for ICH and PHE collinearity, which also showed no significant association between sex or FAST treatment groups with outcome (supplemental table 4).

## DISCUSSION

In this post hoc analysis of the FAST trial in patients with ICH, men had a higher rate of early ICH expansion at 24 hours and a larger absolute early PHE expansion up to 72 hours compared to women. A repeated measures analysis that accounted for the FAST trial treatment assignment showed that the PHE trajectory (PHE volumes across CT brain scans), was significantly higher in men. There was a robust association between PHE expansion and poor outcome, but despite worse imaging characteristics in men, sex was not associated with poor outcome. These findings indicate that PHE, the imaging surrogate of inflammatory secondary injury after ICH, segregates by sex.

### ICH volumes and hematoma expansion

In line with our findings, a post-hoc analysis of the ATACH-2 trial reported similar admission ICH volumes between men and women, and a non-significantly larger proportion of men experienced hematoma expansion at 24 hours.^15^ However, this finding may have been confounded by a higher proportion of deep hematomas among men, as hematoma location has been shown to interact with the ATACH intervention (intensive versus standard blood pressure lowering) with regards to the risk of hematoma expansion.^18^ A post-hoc analysis of the INTERACT trials found larger admission ICH volumes in men, and no differences in the rate of hematoma expansion between men and women, hypothetically influenced by differences in CT timing relative to symptom onset created by segmentation of the randomized data set by sex (INTERACT II included patients who received admission CT >4 hours after symptom onset).^19^ Our study obtained admission CT within 3 hours of symptom onset, found no differences in ICH location or symptom onset to treatment time between the sexes, rigorously adjusted for the FAST-trial treatment, and found a significantly greater rate of hematoma expansion in men that could not be attributed to systolic blood pressure differences and persisted in the subgroup of FAST participants assigned to placebo. This finding suggests that sex-specific factors, e.g., biological or psychosocial, might play a role in the different rates of early hematoma expansion in men and women.^20–22^

### PHE volumes and trajectory

Few data exist on sex differences in PHE trajectories. One retrospective cohort study demonstrated smaller PHE volumes on follow-up imaging in women, but imaging data were pooled from broad time windows and were not correlated with longer-term outcomes.^23^ Our study used more precise imaging timing and blinded volume measurements. Apart from greater lesion volume expansion in men, which we found for both ICH and PHE volumes, the repeated measures analysis demonstrated an overall higher trajectory of PHE volumes in men compared to women, a difference not seen in the ICH trajectory. In contrast to early lesion expansion, which is more susceptible to confounding by CT timing relative to bleed onset, the PHE trajectory up to 72 hours after baseline CT is more likely to represent a finding driven by biological factors as confounding from initial CT timing becomes less impactful over time. A more robust pro-inflammatory response in men may lead to increased PHE formation.^24^ ^25^ As novel targets and therapies of immune-mediated inflammation after ICH are being investigated, the finding of a higher PHE trajectory in men may become important for study planning, and subgroup analyses.^26–28^

### Sex, PHE, and Outcomes

PHE volume is strongly correlated with ICH volume. Early studies, that did not fully address the collinearity between ICH and PHE, found no or only weak associations between PHE volumes and functional outcome, likely due to differences in imaging or volumetric methodology and measurement of absolute or relative (ratio to ICH volume) PHE volumes.^9^ ^29–31^ Our study confirms prior findings of an association between early PHE growth and outcome, with the novel finding that this association persists for PHE expansion that occurs up to 72 hours after the index ICH.^8 9 11 31–34^

We found similar outcomes in men and women, and when adjusting for PHE and ICH volumes and baseline mRS we did not find an independent effect of sex on outcomes. This stands in contrast to a prior study that reported worse outcomes in women.^35^ ^15^ However, men and women in those analyses were not balanced for surgical ICH evacuation, nor did the studies adjust for ICH location or onset to imaging-time.^15^ ^36^ In our study, the absence of an association between female sex and more favorable outcome despite smaller ICH and PHE volumes on follow-up imaging may have several explanations. There may be residual confounding that our multivariate models did not address. Women in the FAST cohort were older than men and had a higher prevalence of premorbid hypertension (both statistically not significant) which might correlate with higher baseline comorbidities in women not recorded in the FAST trial. Women may have received less aggressive care^37^, or may have had a higher incidence of in-hospital complications.^37^ ^38^ Given that an age-dependent increased risk of poor outcomes in women has been reported, the overall younger age of the FAST participants compared to prior cohorts may explain discrepancies with prior studies as well.^39^

Our study may have implications for future research. Due to the higher rate of early lesion expansion in men, prevention of hematoma or PHE expansion may yield greater outcome benefit in men. This might influence study design, patient recruitment strategies, and pre-specification of subgroup analyses in future interventional trials. Our findings also suggest that focusing on sex-specific factors that contribute to PHE, a major cause of secondary injury after ICH, may allow novel mechanistic insight into PHE. And lastly, our study highlights a need to identify social factors, such as less effective prevention or impeded access to health care that may contribute to equally poor outcome in women despite more favorable ICH and PHE characteristics in women.

### Limitations

This study has limitations. First, this was an observational analysis that cannot infer causality. Second, volumetric analysis of PHE based on hypoattenuating tissue on CT may be suboptimal as gliosis, chronic microangiopathic changes or periventricular transependymal flow with hydrocephalus may be confounders.^40^ However, measurement accuracy would not be expected to vary by sex. Third, ideally volumetric analyses should be adjusted for whole brain volumes, but these data were not at our disposal. Fourth, the FAST imaging protocol captured neuroimaging changes only up to 72 hours and thus did not capture the complete trajectory of PHE. Furthermore, we did not have data available on pre-admission hypertension severity and medication use to provide insight into underlying vascular health, information on antiplatelet use before the index ICH, or information on the specific etiologies of ICH comparing men and women. Lastly, the selective inclusion criteria of the FAST trial may have skewed our results. By design the trial excluded patients with longer time intervals from symptom onset to hospital admission which may have minimized differences in social support and access to care that might exist in broader populations of patients with ICH and might influence outcomes.^35^ The lower proportion of enrolled women, while possibly attributable to a lower incidence of ICH in women, may have been influenced by selection bias and may thus have impacted our findings.

In conclusion, we found that PHE volume trajectories after spontaneous ICH segregated by sex, with a more pronounced upward trajectory of PHE volumes in men compared to women. While PHE expansion was associated with worse outcomes independent of ICH volumes, there was no effect of sex on outcomes. Women had equally poor outcomes as men, despite less PHE and ICH expansion in women. Mechanisms explaining differences in PHE trajectories, despite equally poor functional outcomes between men and women, remain to be elucidated.

## Data Availability

Anonymized data from the FAST trial (Factor VII for Acute Hemorrhagic Stroke) can be requested through the Virtual International Stroke Trials Archive (VISTA, www.virtualtrialsarchive.org).

## Acknowledgements

A summary of this work was presented as an oral platform presentation at the International Stroke Conference 2023, Dallas, USA.

## Competing Interests

Dr. Witsch: fees for medicolegal consulting, grant funding from the American Heart Association (23CDA1053561). Dr. Cao: no disclosures. Dr. Song: no disclosures. Y.Luo: no disclosures. Dr. Sloane: no disclosures. Dr. Rothstein: no disclosures. Dr. Favilla: no disclosures. Dr. Cucchiara: Honoraria; Modest; Astra Zeneca, Other; Modest; Bayer. Dr. Kasner: Other; Modest; Bayer, Bristol Meyers Squibb, Boehringer Ingelheim, Medtronic, Diamedica, Research Grant; Modest; Genentech, WL Gore. Dr. Messé: no disclosures. Dr. Choi: no disclosures. Dr. McCullough: NIH/NINDS (RF1AG058463, R01NS103592, R01NS094543). Dr. Mayer: no disclosures. Dr. Gusdon: NINDS (1K23NS121628-01A1).

## Funding

There was no targeted funding for this study.

## Notes

### Competing Interest Statement

Dr. Witsch: fees for medicolegal consulting, royalties from a Springer textbook on Stroke, and grant funding from the American Heart Association Career Development Award (https://doi.org/10.58275/AHA.23CDA1053561.pc.gr.168057). Dr. Cao: no disclosures. Dr. Song: grant funding from the American Heart Association Career Development Award (938082). Y.Luo: no disclosures. Dr. Sloane: no disclosures. Dr. Rothstein: no disclosures. Dr. Favilla: no disclosures. Dr. Cucchiara: Honoraria; Modest; Astra Zeneca, Other; Modest; Bayer. Dr. Kasner: Other; Modest; Bayer, Bristol Meyers Squibb, Boehringer Ingelheim, Medtronic, Diamedica, Research Grant; Modest; Genentech, WL Gore. Dr. Messe: no disclosures. Dr. Choi: no disclosures. Dr. McCullough: NIH/NINDS (RF1AG058463, R01NS103592, R01NS094543). Dr. Mayer: no disclosures. Dr. Gusdon: NINDS (1K23NS121628-01A1).

### Author Declarations

This study is a secondary analysis of data previously collected and was exempt from additional institutional review board approval. Exempt from full board review was granted by the Institutional Review Board of the University of Pennsylvania.

## REFERENCES

1. Greenberg SM, Ziai WC, Cordonnier C, et al. 2022 Guideline for the Management of Patients With Spontaneous Intracerebral Hemorrhage: A Guideline From the American Heart Association/American Stroke Association. Stroke 2022:101161STR0000000000000407. doi: 10.1161/STR.0000000000000407 [published Online First: 20220517]

2. van Asch CJ, Luitse MJ, Rinkel GJ, et al. Incidence, case fatality, and functional outcome of intracerebral haemorrhage over time, according to age, sex, and ethnic origin: a systematic review and meta-analysis. Lancet Neurol 2010;9(2):167–76. doi: 10.1016/S1474-4422(09)70340-0 [published Online First: 20100105]

3. Dowlatshahi D, Demchuk AM, Flaherty ML, et al. Defining hematoma expansion in intracerebral hemorrhage: relationship with patient outcomes. Neurology 2011;76(14):1238–44. doi: 10.1212/WNL.0b013e3182143317 [published Online First: 2011/02/25]

4. Venkatasubramanian C, Kleinman JT, Fischbein NJ, et al. Natural history and prognostic value of corticospinal tract Wallerian degeneration in intracerebral hemorrhage. J Am Heart Assoc 2013;2(4):e000090. doi: 10.1161/jaha.113.000090 [published Online First: 2013/08/06]

5. Staykov D, Wagner I, Volbers B, et al. Natural course of perihemorrhagic edema after intracerebral hemorrhage. Stroke 2011;42(9):2625–9. doi: 10.1161/STROKEAHA.111.618611 [published Online First: 20110707]

6. Ironside N, Chen CJ, Ding D, et al. Perihematomal Edema After Spontaneous Intracerebral Hemorrhage. Stroke 2019;50(6):1626–33. doi: 10.1161/STROKEAHA.119.024965 [published Online First: 20190502]

7. Lee KH, Lioutas VA, Marchina S, et al. The Prognostic Roles of Perihematomal Edema and Ventricular Size in Patients with Intracerebral Hemorrhage. Neurocrit Care 2022 doi: 10.1007/s12028-022-01532-0 [published Online First: 20220608]

8. Urday S, Beslow LA, Dai F, et al. Rate of Perihematomal Edema Expansion Predicts Outcome After Intracerebral Hemorrhage. Crit Care Med 2016;44(4):790–7. doi: 10.1097/ccm.0000000000001553 [published Online First: 2016/01/13]

9. Murthy SB, Moradiya Y, Dawson J, et al. Perihematomal Edema and Functional Outcomes in Intracerebral Hemorrhage: Influence of Hematoma Volume and Location. Stroke 2015;46(11):3088–92. doi: 10.1161/strokeaha.115.010054 [published Online First: 2015/09/24]

10. Marchina S, Trevino-Calderon JA, Hassani S, et al. Perihematomal Edema and Clinical Outcome After Intracerebral Hemorrhage: A Systematic Review and Meta-Analysis. Neurocrit Care 2022;37(1):351–62. doi: 10.1007/s12028-022-01512-4 [published Online First: 20220516]

11. Grunwald Z, Beslow LA, Urday S, et al. Perihematomal Edema Expansion Rates and Patient Outcomes in Deep and Lobar Intracerebral Hemorrhage. Neurocrit Care 2017;26(2):205–12. doi: 10.1007/s12028-016-0321-3 [published Online First: 2016/11/16]

12. Gokhale S, Caplan LR, James ML. Sex differences in incidence, pathophysiology, and outcome of primary intracerebral hemorrhage. Stroke 2015;46(3):886–92. doi: 10.1161/STROKEAHA.114.007682 [published Online First: 2015/02/07]

13. Roquer J, Rodriguez-Campello A, Jimenez-Conde J, et al. Sex-related differences in primary intracerebral hemorrhage. Neurology 2016;87(3):257–62. doi: 10.1212/wnl.0000000000002792 [published Online First: 2016/06/10]

14. Falcone GJ, Biffi A, Brouwers HB, et al. Predictors of hematoma volume in deep and lobar supratentorial intracerebral hemorrhage. JAMA Neurol 2013;70(8):988–94. doi: 10.1001/jamaneurol.2013.98

15. Fukuda-Doi M, Yamamoto H, Koga M, et al. Sex Differences in Blood Pressure-Lowering Therapy and Outcomes Following Intracerebral Hemorrhage: Results From ATACH-2. Stroke 2020;51(8):2282–86. doi: 10.1161/STROKEAHA.120.029770 [published Online First: 20200706]

16. Mayer SA, Brun NC, Begtrup K, et al. Efficacy and safety of recombinant activated factor VII for acute intracerebral hemorrhage. N Engl J Med 2008;358(20):2127–37. doi: 10.1056/NEJMoa0707534 [published Online First: 2008/05/16]

17. Kuohn LR, Witsch J, Steiner T, et al. Early Deterioration, Hematoma Expansion, and Outcomes in Deep Versus Lobar Intracerebral Hemorrhage: The FAST Trial. Stroke 2022;53(8):2441–48. doi: 10.1161/STROKEAHA.121.037974 [published Online First: 20220401]

18. Leasure AC, Qureshi AI, Murthy SB, et al. Association of Intensive Blood Pressure Reduction With Risk of Hematoma Expansion in Patients With Deep Intracerebral Hemorrhage. JAMA Neurol 2019 doi: 10.1001/jamaneurol.2019.1141 [published Online First: 2019/05/14]

19. Sandset EC, Wang X, Carcel C, et al. Sex differences in treatment, radiological features and outcome after intracerebral haemorrhage: Pooled analysis of Intensive Blood Pressure Reduction in Acute Cerebral Haemorrhage trials 1 and 2. Eur Stroke J 2020;5(4):345–50. doi: 10.1177/2396987320957513 [published Online First: 20200920]

20. Bushnell C, McCullough LD, Awad IA, et al. Guidelines for the prevention of stroke in women: a statement for healthcare professionals from the American Heart Association/American Stroke Association. Stroke 2014;45(5):1545–88. doi: 10.1161/01.str.0000442009.06663.48 [published Online First: 20140206]

21. Zelniker TA, Ardissino M, Andreotti F, et al. Comparison of the Efficacy and Safety Outcomes of Edoxaban in 8040 Women Versus 13 065 Men With Atrial Fibrillation in the ENGAGE AF-TIMI 48 Trial. Circulation 2021;143(7):673–84. doi: 10.1161/CIRCULATIONAHA.120.052216 [published Online First: 20210215]

22. Abou-Ismail MY, Citla Sridhar D, Nayak L. Estrogen and thrombosis: A bench to bedside review. Thromb Res 2020;192:40–51. doi: 10.1016/j.thromres.2020.05.008 [published Online First: 20200511]

23. Wagner I, Volbers B, Kloska S, et al. Sex differences in perihemorrhagic edema evolution after spontaneous intracerebral hemorrhage. Eur J Neurol 2012;19(11):1477–81. doi: 10.1111/j.1468-1331.2011.03628.x [published Online First: 2012/01/10]

24. Gupta S, Nakabo S, Blanco LP, et al. Sex differences in neutrophil biology modulate response to type I interferons and immunometabolism. Proc Natl Acad Sci U S A 2020;117(28):16481–91. doi: 10.1073/pnas.2003603117 [published Online First: 20200629]

25. Ahnstedt H, Patrizz A, Roy-O’Reilly M, et al. Abstract TMP36: Sex Differences in Neutrophil-T Cell Immune Responses and Outcome After Ischemic Stroke in Aged Mice. Stroke 2018;49(Suppl_1):ATMP36-ATMP36. doi: doi:10.1161/str.49.suppl_1.TMP36

26. Ohashi SN, DeLong JH, Kozberg MG, et al. Role of Inflammatory Processes in Hemorrhagic Stroke. Stroke 2023;54(2):605–19. doi: 10.1161/STROKEAHA.122.037155 [published Online First: 20230105]

27. Ngo ATP, Gollomp K. Building a better NET: Neutrophil extracellular trap targeted therapeutics in the treatment of infectious and inflammatory disorders. Research and Practice in Thrombosis and Haemostasis 2022;6(7):e12808. doi: 10.1002/rth2.12808

28. Banerjee A, Wang J, Bodhankar S, et al. Phenotypic changes in immune cell subsets reflect increased infarct volume in male vs. female mice. Translational stroke research 2013;4(5):554–63. doi: 10.1007/s12975-013-0268-z

29. Arima H, Wang JG, Huang Y, et al. Significance of perihematomal edema in acute intracerebral hemorrhage: the INTERACT trial. Neurology 2009;73(23):1963–8. doi: 10.1212/WNL.0b013e3181c55ed3 [published Online First: 2009/12/10]

30. Appelboom G, Bruce SS, Hickman ZL, et al. Volume-dependent effect of perihaematomal oedema on outcome for spontaneous intracerebral haemorrhages. J Neurol Neurosurg Psychiatry 2013;84(5):488–93. doi: 10.1136/jnnp-2012-303160 [published Online First: 20130123]

31. Yang J, Arima H, Wu G, et al. Prognostic significance of perihematomal edema in acute intracerebral hemorrhage: pooled analysis from the intensive blood pressure reduction in acute cerebral hemorrhage trial studies. Stroke 2015;46(4):1009–13. doi: 10.1161/strokeaha.114.007154 [published Online First: 2015/02/26]

32. Wu TY, Sharma G, Strbian D, et al. Natural History of Perihematomal Edema and Impact on Outcome After Intracerebral Hemorrhage. Stroke 2017;48(4):873–79. doi: 10.1161/STROKEAHA.116.014416

33. Murthy SB, Urday S, Beslow LA, et al. Rate of perihaematomal oedema expansion is associated with poor clinical outcomes in intracerebral haemorrhage. J Neurol Neurosurg Psychiatry 2016;87(11):1169–73. doi: 10.1136/jnnp-2016-313653

34. Li N, Liu YF, Ma L, et al. Association of molecular markers with perihematomal edema and clinical outcome in intracerebral hemorrhage. Stroke 2013;44(3):658–63. doi: 10.1161/strokeaha.112.673590 [published Online First: 2013/02/09]

35. Bueno Alves M, Freitas de Carvalho JJ, Álvares Andrade Viana G, et al. Gender Differences in Patients with Intracerebral Hemorrhage: A Hospital-Based Multicenter Prospective Study. Cerebrovascular diseases extra 2012;2(1):63–70. doi: 10.1159/000343187

36. Xia F, Chen Y, Hu X. Letter by Xia, et al Regarding Article, “Sex Differences in Blood Pressure-Lowering Therapy and Outcomes Following Intracerebral Hemorrhage: Results From ATACH-2”. Stroke 2020;51(12):e359. doi: 10.1161/STROKEAHA.120.031689 [published Online First: 20201123]

37. Wang SS, Bogli SY, Nierobisch N, et al. Sex-Related Differences in Patients’ Characteristics, Provided Care, and Outcomes Following Spontaneous Intracerebral Hemorrhage. Neurocrit Care 2022;37(1):111–20. doi: 10.1007/s12028-022-01453-y [published Online First: 20220407]

38. Otite FO, Khandelwal P, Malik AM, et al. Ten-Year Temporal Trends in Medical Complications After Acute Intracerebral Hemorrhage in the United States. Stroke 2017;48(3):596–603. doi: 10.1161/STROKEAHA.116.015746 [published Online First: 20170222]

39. Umeano O, Phillips-Bute B, Hailey CE, et al. Gender and age interact to affect early outcome after intracerebral hemorrhage. PloS one 2013;8(11):e81664. doi: 10.1371/journal.pone.0081664 [published Online First: 20131127]

40. Carhuapoma JR, Hanley DF, Banerjee M, et al. Brain edema after human cerebral hemorrhage: a magnetic resonance imaging volumetric analysis. Journal of neurosurgical anesthesiology 2003;15(3):230–3. doi: 10.1097/00008506-200307000-00010 [published Online First: 2003/06/27]

