## supplemental for "Sex differences in perihematomal edema volume and outcome after intracerebral hemorrhage"

**Supplemental Methods**

**Supplemental Figure 1-4**

**Supplemental Table 1-5**

**Supplemental Methods**

Design and Participants

Patients were eligible for enrollment if they were 18 years of age or older and had a spontaneous ICH documented on a brain CT within 3 hours of symptoms onset. Patients were excluded from FAST if any of the following characteristics were present: admission GCS of 5 or less, surgical ICH evacuation planned within 24 hours of admission, secondary ICH due to vascular malformation or trauma; known anticoagulant use, thrombocytopenia, or history of coagulopathy, sepsis, crush injury, disseminated intravascular coagulation, pregnancy, prior disability (mRS > 2), known thrombotic or vaso-occlusive disease (includes deep vein thrombosis, myocardial and cerebral infarction) within 30 days before ICH onset. A total of 841 patients were enrolled between May 2005 and February 2007 at 122 sites across 22 countries. Enrolled patients were randomly assigned to receive one of the three treatments within 4 hours of stroke onset: placebo, 20 µg of rFVIIa per kilogram body weight, or 80 µg of rFVIIa per kilogram body weight. In this analysis, we excluded patients with infratentorial hemorrhage (n=40), due to the limitations of CT-based assessment of PHE in the posterior fossa.

Written informed consent for trial participation was provided by each patient or their legally authorized representative. Institutional review board approval was obtained at each enrolling site of the FAST trial. Given that only deidentified, publicly available data were used, this retrospective study was exempt from additional, local IRB approval.

Statistical Methods

Statistical analyses were performed using SPSS (version 29, IBM, Armonk, New York) and R (version 4.2.2, R Foundation for Statistical Computing). Descriptive statistics are presented as counts (percent) and mean (standard deviation [SD]) or median (interquartile range [IQR]) for discrete and continuous variables, respectively. For unadjusted comparisons of discrete variables, we used chi-squared testing. For comparisons of lesion volumes, we used non-parametric statistical tests: Mann Whitney U tests for between-group comparisons (e.g., lesion volumes between men and women), and the Wilcoxon signed rank test for within-group comparisons (e.g., lesion volumes between two timepoints).

**Supplemental Figure 1.** STROBE flowchart of included and excluded patients

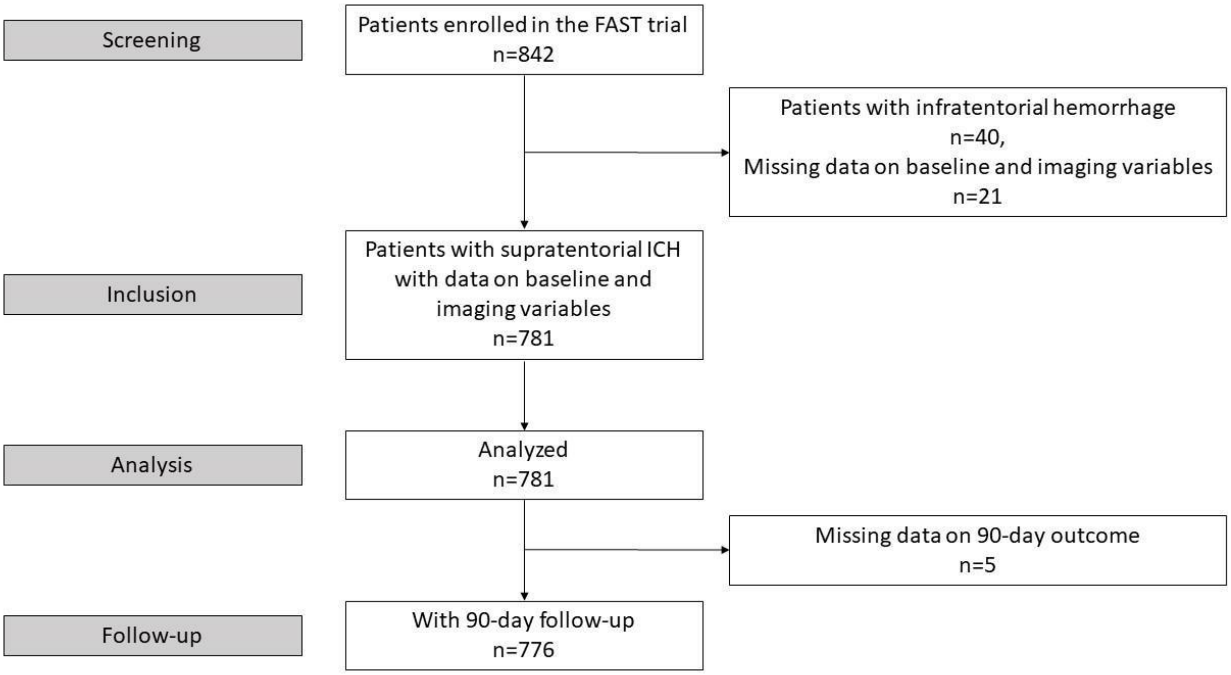

**Supplemental Figure 2.** Distribution of ICH volumes

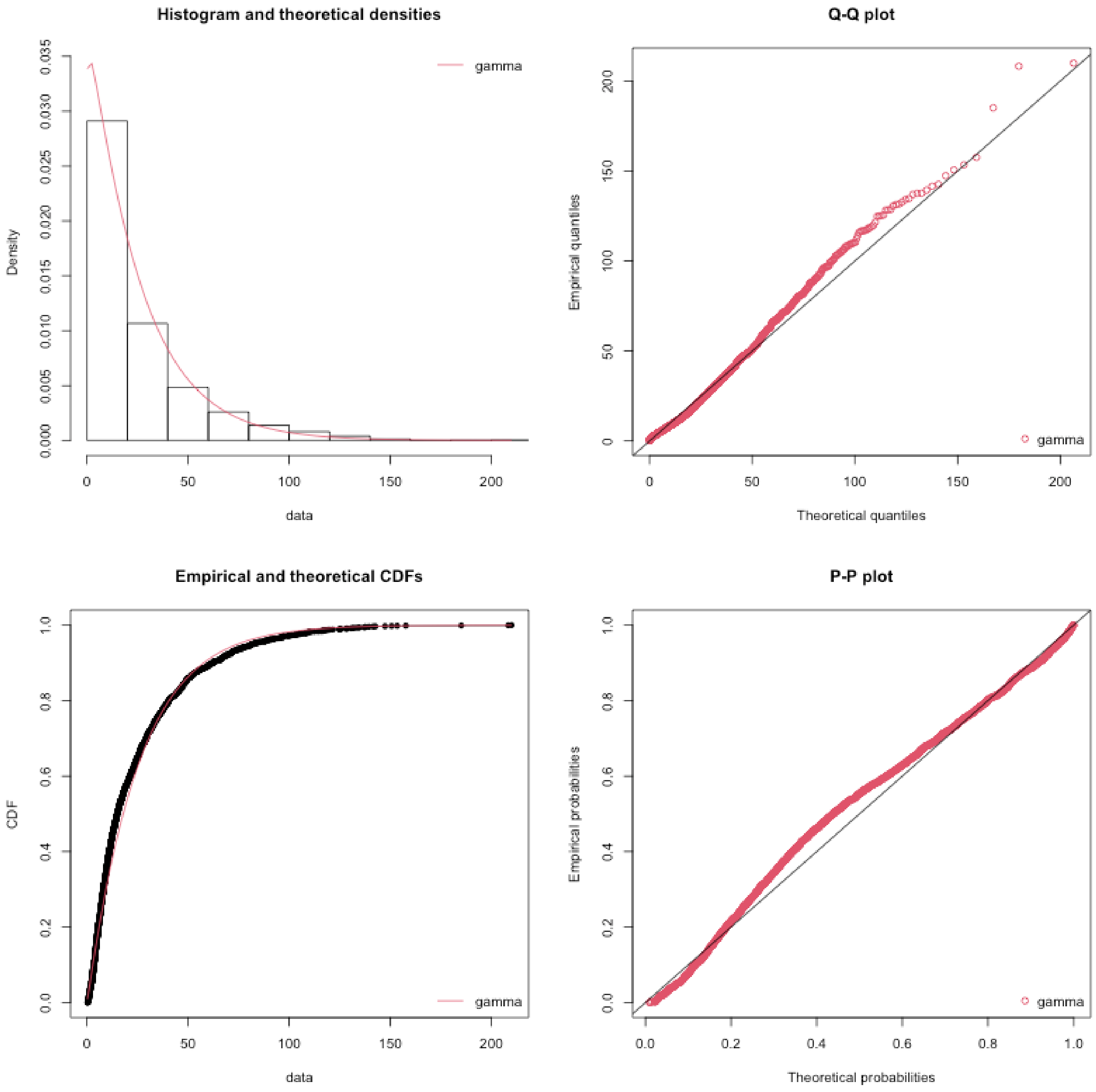


**Assessing the distribution of ICH volume data**.
Top-left: Histogram of the data and theoretical densities; Top-right: Quantile-Quantile (Q-Q) plot; Bottom left: Empirical and theoretical cumulative distribution function, probability-probability (P-P) plot. CDF, cumulative distribution function.

**Supplemental Figure 3.** Distribution of PHE volumes

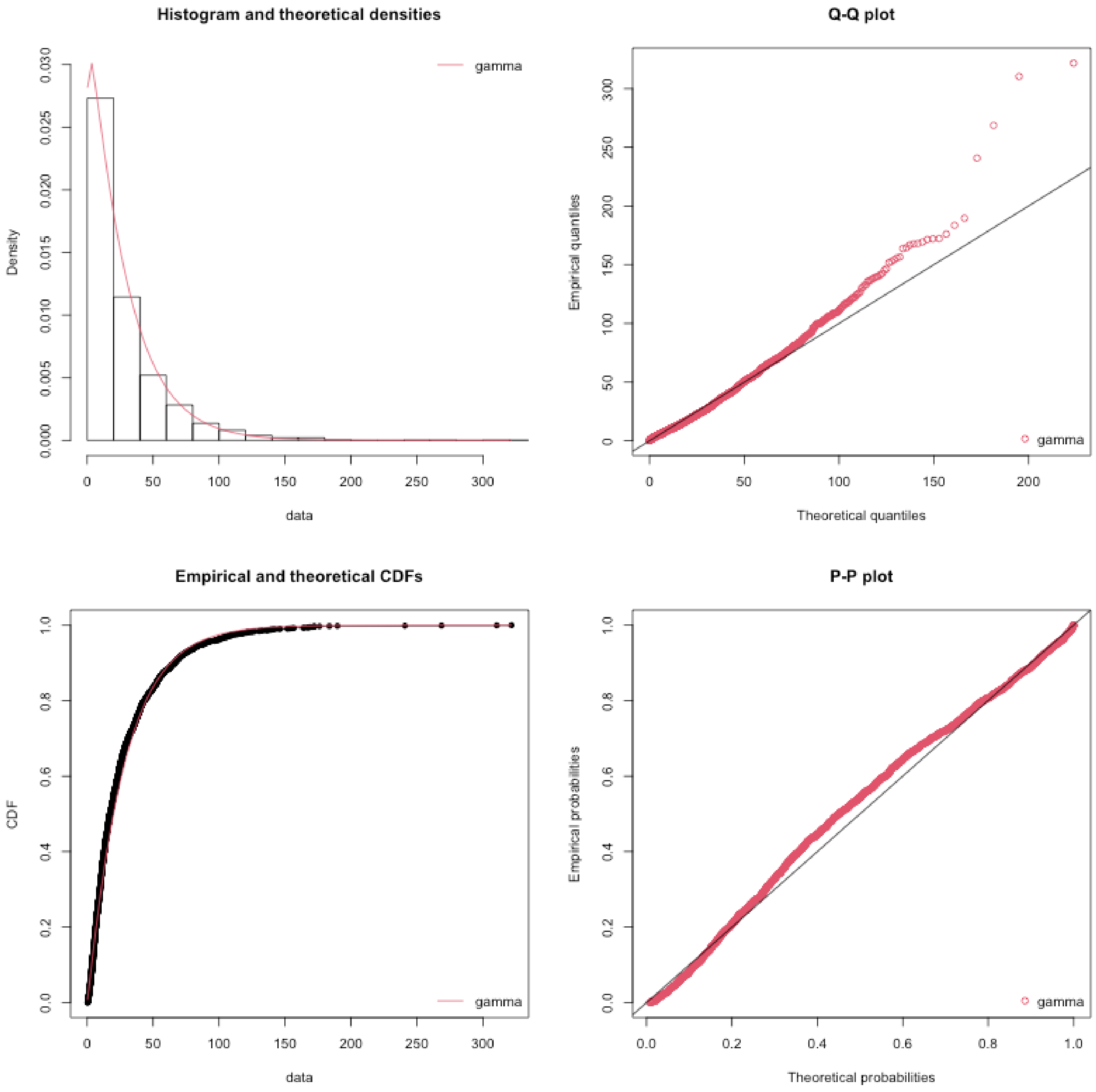


**Assessing the distribution of PHE volume data**.
Top-left: Histogram of the data and theoretical densities; Top-right: Q-Q plot; Bottom left: Empirical and theoretical cumulative distribution function, P-P plot.

**Supplemental Figure 4.** Distribution of total lesion volumes

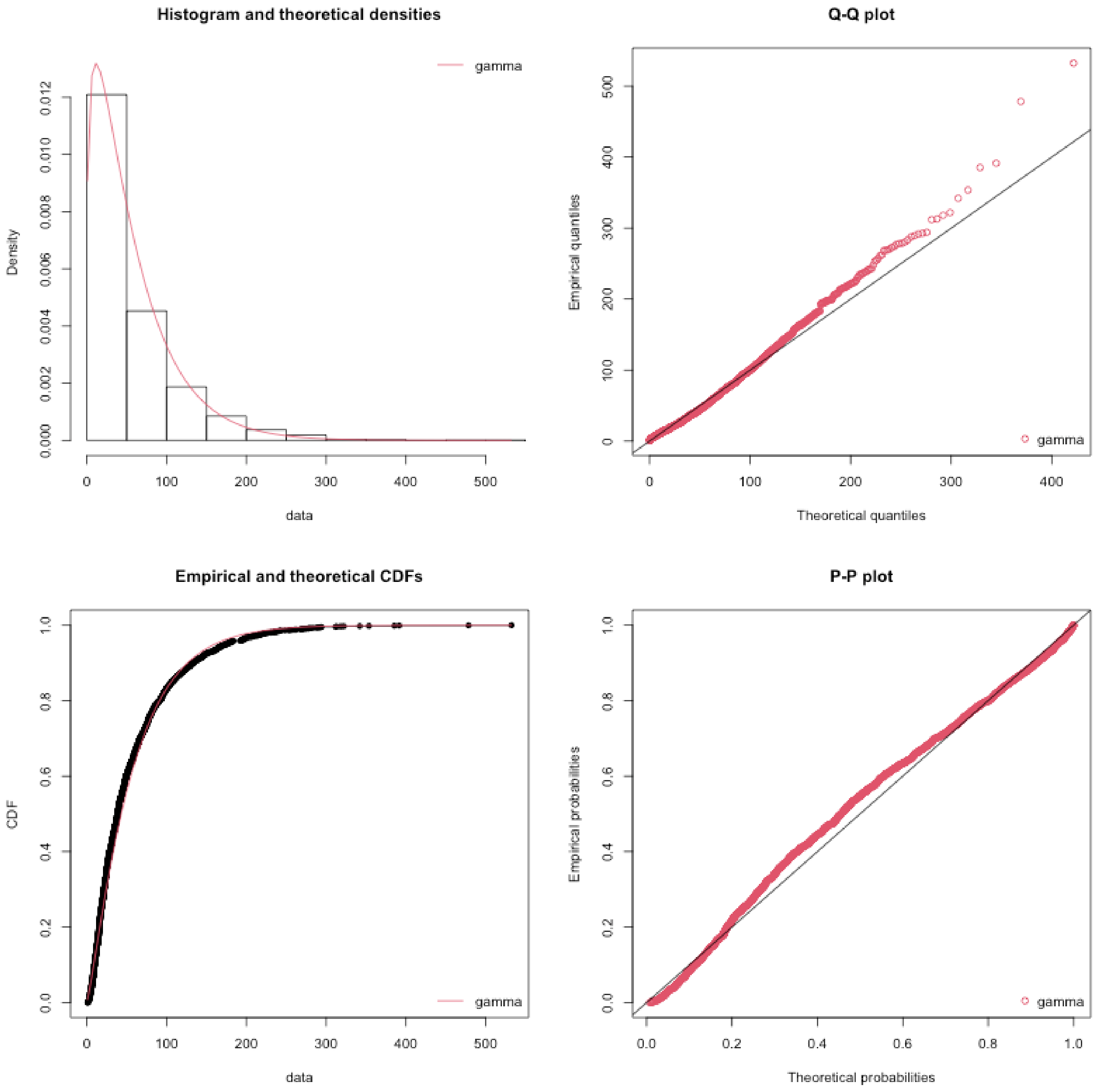


**Assessing the distribution of total lesion volume data**.

Top-left: Histogram of the data and theoretical densities; Top-right: Q-Q plot; Bottom left: Empirical and theoretical cumulative distribution function, P-P plot.

| Supplemental Table 1. Systolic Blood Pressure in the FAST Cohort*, Stratified by Sex | | | |
|---|---|---|---|
| | **Men** | **Women** | **p** |
| Systolic Blood Pressure [mmHg] on Admission, mean (SD) | 178.82 (29.6) | 183.07 (30.4) | 0.054 |
| Systolic Blood Pressure [mmHg] Visit #2, mean (SD) | 173.48 (26.8) | 173.27 (26.1) | 0.9 |
| Systolic Blood Pressure [mmHg] Visit #3, mean (SD) | 167.26 (26.2) | 165.23 (26.3) | 0.3 |
| Systolic Blood Pressure [mmHg] Visit #4, mean (SD) | 155.17 (23.6) | 154.53 (24.6) | 0.7 |

*Participants with infratentorial hemorrhages (n=40) were excluded from this post hoc analysis

SD, standard deviation

Visit #2-4: at 1 hour, at 24 hours after drug administration, and on days 2 and 3.

| Supplemental Table 2. Hematoma Expansion in the FAST Treatment Groups*, Stratified by Sex | | | |
|---|---|---|---|
| | **Men, Hematoma Expansion** | **Women, Hematoma Expansion** | **p** |
| Placebo, n=246 (160 men, 86 women) | 61 (38) | 22 (26) | 0.047 |
| Low dose, n=257 (155 men, 102 women) | 51 (33) | 25 (25) | 0.15 |
| High dose, n=278 (170 men, 108 women) | 48 (28) | 17 (16) | 0.016 |

*Participants with infratentorial hemorrhages (n=40) were excluded from this post hoc analysis

Hematoma expansion defined as hematoma volume increase of ≥33% or 6 mL at 24 hours

Low dose, 20 µg of rFVIIa per kilogram body weight
High dose, 80 µg of rFVIIa per kilogram body weight

Data are represented as numbers (%)

**Supplemental Table 3. Two-way ANOVA testing for independence of lesion volumes of sex**

| CT timepoints | p |
|---|---|
| Baseline | 0.33 |
| 24 hours | 0.82 |
| 72 hours | 0.74 |

**Supplemental Table 4. Logistic Mixed Model of Poor Outcome (mRS 4-6)**

| | PHE model | | ICH model | |
|---|---|---|---|---|
| **Variable** | **OR (CI)** | **p** | **OR (CI)** | **p** |
| **Lesion volume*** | 2.01 (1.47-2.70) | <0.001 | 21.1 (4.65-99.6) | <0.001 |
| **Female sex** | 0.70 (0.04-12.3) | 0.80 | 0.21 (0.00004-1033) | 0.72 |
| **Interaction: lesion volume x female sex** | 1.02 (0.92-1.13) | 0.70 | NA# | NA# |
| **Timepoint 24 hours** | 0.30 (0.08-1.04) | 0.056 | 0.30 (0.007-12.7) | 0.52 |
| **Timepoint 72 hours** | 0.11 (0.03-0.50) | 0.004 | 0.45 (0.01-19.9) | 0.68 |
| **Age** | 1.31 (1.20-1.43) | <0.001 | 1.75 (1.22-2.51) | 0.002 |
| **Admission GCS** | 0.20 (0.11-0.35) | <0.001 | 0.03 (0.003-0.28) | 0.002 |
| **Lobar hematoma location** | 0.31 (0.01-6.42) | 0.45 | 4.129e-7 (5.661e-13-0.30) | 0.03 |
| **Presence of IVH** | 11.13 (1.34-93.7) | 0.03 | 54.6 (0.009-31,8061) | 0.37 |
| **rFVIIa (low dose)^** | 1.45 (0.12-17.1) | 0.77 | 2.03 (0.0001-41,357) | 0.89 |
| **rFVIIa (high dose)^** | 2.83 (0.26-31.2) | 0.40 | 0.55 (2.953e-5-10,301) | 0.91 |
| **Baseline mRS 1-2** | 45.2 (2.39-854.1) | 0.01 | 798,109 (2.27-) | 0.04 |

Abbreviations: OR, odds ratio; CI, 95% confidence interval; GCS, Glasgow Coma Scale; ICH, intracerebral hemorrhage; IVH, intraventricular hemorrhage; PHE, perihematomal edema; rFVIIa, recombinant factor VIIa; mRS, modified Rankin Scale.
*Refers to PHE lesion volume in the PHE model and ICH lesion volume in the ICH model, ORs represent per 5 ml increase in lesion volume
#The ICH model did not allow for the inclusion of the interaction variable
^Compared to the FAST placebo arm as a reference

**Supplemental Table 5. PHE-Residual Logistic Mixed Model of Poor Outcome (mRS 4-6) at 90 Days**

| Variable | Beta (standard error) | p |
|---|---|---|
| Residual from PHE on ICH linear regression | 0.003 (0.03) | 0.92 |
| Female sex | -1.02 (2.2) | 0.64 |
| Timepoint 24 hours | 0.062 (0.90) | 0.94 |
| Timepoint 72 hours | -0.028 (1.07) | 0.98 |
| Age | 0.49 (0.10) | <0.001 |
| Admission GCS | -3.40 (0.62) | <0.001 |
| Lobar hematoma location | 2.57 (2.94) | 0.38 |
| Presence of IVH | 5.77 (2.28) | 0.01 |
| rFVIIa (low dose) | -0.38 (2.60) | 0.88 |
| rFVIIa (high dose) | -0.40 (2.55) | 0.88 |
| Baseline mRS 1-2 | 7.01 (3.24) | 0.03 |

Abbreviations: GCS, Glasgow Coma Scale; ICH, intracerebral hemorrhage; IVH, intraventricular hemorrhage; PHE, perihematomal edema; rFVIIa, recombinant factor VIIa; mRS, modified Rankin Scale.
